# Mental health patterns and associated social determinants among university and college students in Sub-Saharan Africa during the COVID-19 pandemic era: A Scoping Review Protocol

**DOI:** 10.1101/2024.10.08.24315086

**Authors:** Memory Muturiki, Nontsikelelo Mapukata, Lawrence Chauke, Sara Jewett

## Abstract

**Objective:** The objective of this scoping review is to identify and map the literature that documents student mental health patterns and associated social determinants during the COVID-19 pandemic era in Sub-Saharan Africa (SSA) universities and colleges.

**Introduction:** The rationale for the scoping review is to identify gaps in existing literature. Most of the current data on student mental health and the prevalence of mental disorders in universities and colleges during the COVID-19 years is from the global north. There is limited data for Sub-Saharan African universities on student mental health and the extent to which social determinants contribute. The review will also provide areas for further research among Higher Education Institutions (HEIs) in the SSA region.

**Eligibility criteria:** Only documents published during the years 2020 to 2023 from SSA will be included and documents from outside the period and outside the geographic region will not be considered. HEIs will be post-school education universities and colleges, and so school level reports will be excluded.

**Methods:** This scoping review will be conducted using various search engines on existing data for student mental health and social determinants in the era of COVID-19 (years 2020 to 2023). Search engines will include MEDLINE (PubMed), PsycInfo, EMBASE (Ovid), African Index Medicus, Open Access Journals, CINAHL, JBI Library, Google Scholar, the Cochrane Library (Ovid), and Grey Literature (various relevant health related websites). Search limits will only consider documents written in English language. Search terms will be student mental health, university mental health services, social determinants of health, social determinants of mental health and Sub-Saharan Africa. Documents searched will be uploaded into EndNote 21 and will be coded, and themes will be generated using NVivo-12.

**Results:** Results will be reported using the Scoping Reviews (PRISMA-ScR) guidelines (Wits-JBI evidence synthesis procedure) and presented as tables and graphs. The key findings of the scoping review in together with the research question will be reported once completed.

**Conclusions:** A conclusion based on the scoping review findings along with the objectives will be provided when the review is completed. Main implications of the findings (if any) will also be conveyed.

## Introduction

### Student Mental Health

According to the World Health Organisation (WHO), mental health includes holistic wellbeing, psychological, and social well-being, in a way which enables individuals to cope with stress and manage relationships [1]. Student mental health is the state of mental wellbeing that enables students to cope with the stresses of life and focus on their academic activities [1]. Academic stress is described as the physical and emotional response to the demand of academic activities that is beyond the coping skills of students [2]. Enrolling to study at university or college is a big step which often can be demanding and stressful, additionally students in South Africa face many unique challenges given the historic inequalities of the country [3]. This is also true for students in many resource-limited universities in other lower income countries and in SSA [4]. These challenges include among others, being the first generation in a family to attend university [3, 5], financial problems, food security [6], academic workload, coming from previously disadvantaged areas, rural, and informal settlements [7, 8]. [9] reported on impact of mental disorders and subsequent high-cost outcomes on students and their families because the consequences of academic failure which result in delayed progression, or attrition.

### Student mental health and social determinants

Systematic reviews have pointed to the rising mental disorders among students globally [10]. What is not clearly known is how social determinants of health (SDOH) influence student mental health and why some students seem are more at risk of developing mental disorders than others. SDOH include the environments where people are born, live, study, and perform their daily activities [11]. Social determinants of mental health include some behaviours [12], such as smoking cigarettes, use of drugs as cannabis and drinking alcohol [13].

### Student mental health and COVID-19

The WHO estimated the prevalence of mental illness in young adults is 20-25% [14] and COVID-19 is known to have pushed that up to close to 30% [15]. Given the impact of the COVID-19 pandemic on mental health services in the public health system [16] and on student mental health [17] there are important considerations that universities should consider in designing support for the more vulnerable students, such as the role of social determinants among students [6]. In addition, the COVID-19 brought the mental health struggles of students to surface because of the direct impact of lockdowns on education including the shift from traditional teaching and learning methodologies, the sudden migration to online learning platforms and the social distancing protocols [17, 18].

An initial search of PubMed and PsycInfo will be performed to check if there are any current existing gaps on the topic for data in SSA universities on student mental health and the extent to which social determinants contribute. This will also assist in informing future research in student mental health among universities in SSA region.

### Objective

The objective of this scoping review is to explore and map the available literature on mental health patterns and associated social determinants among university students in SSA Higher Education Institutions (HEIs) during the COVID-19 pandemic era.

### Rationale for the scoping review

The justification for this is because most of the literature on student mental health is from the Global North. The existing gap is lack of research from SSA and given the existing socio-political and economic challenges in SSA [4]. There are many studies and systematic reviews published on student mental health during COVID-19 and so a scoping review is one of the best ways to identify knowledge gaps because scoping reviews a useful approach for reviewing evidence rapidly in emerging fields [19]. Additionally, this scoping review will identify knowledge gaps on the association between social determinants and student mental health in SSA universities and colleges. Results of the scoping review will guide recommendations for the identification of future research initiatives and to enhance the design of student-centred mental health support to be more effective for the more at-risk college students.

## Materials and Methods

This scoping review will use the Wits-JBI methodology for scoping reviews [19]. The protocol title is registered on OSF and can be accessed on https://doi.org/10.17605/OSF.IO/AXK4Y.

### Scoping review question

Is there an existing gap in research and literature on mental health and associated social determinants for HEIs students in limited resource setting such as the SSA?

### Participants

Student participants must be registered in an HEI to be included in this review.

### Search Strategy

The search strategy will attempt to find both published and unpublished relevant studies in consultation with a librarian. The strategy will be initiated by conducting a limited search of MEDLINE (PubMed) and PsycInfo to identify relevant documents (Table 1). The text words in the titles and abstracts of the documents will be used, and the index terms to develop a full search strategy (Table 2). The search strategy will be adapted for each included database in line with Wits- JBI evidence synthesis. The reference list of all included sources of data will be screened for additional studies. Studies published only in English language and published from SSA since the COVID-19 period of 2020 -2023 will be included.

**Table 1.** Initial Search Strategy.

| Concept | Context |
| --- | --- |
| Student mental health<br>or<br>Mental health<br>or<br>Mental disorders<br>or<br>Mental illness<br>or<br>Psychiatric disorders<br>or<br>Mental wellbeing<br>or<br>Student wellness<br>or<br>Social determinants<br>or<br>Social determinants of health<br>or<br>Social determinants of mental health<br>or<br>COVID-19 era<br>or<br>COVID-19 pandemic<br>or<br>COVID-19 outbreak period | University healthcare services<br>or<br>University health services<br>or<br>College health services<br>or<br>University mental health services<br>or<br>Campus-based health services<br>or<br>Sub-Saharan Africa universities<br>or<br>Campus-based clinics<br>or<br>College students<br>or<br>University students<br>or<br>Sub-Saharan Africa Higher Education Institutions<br>or<br>Africa |

**Table 2.**
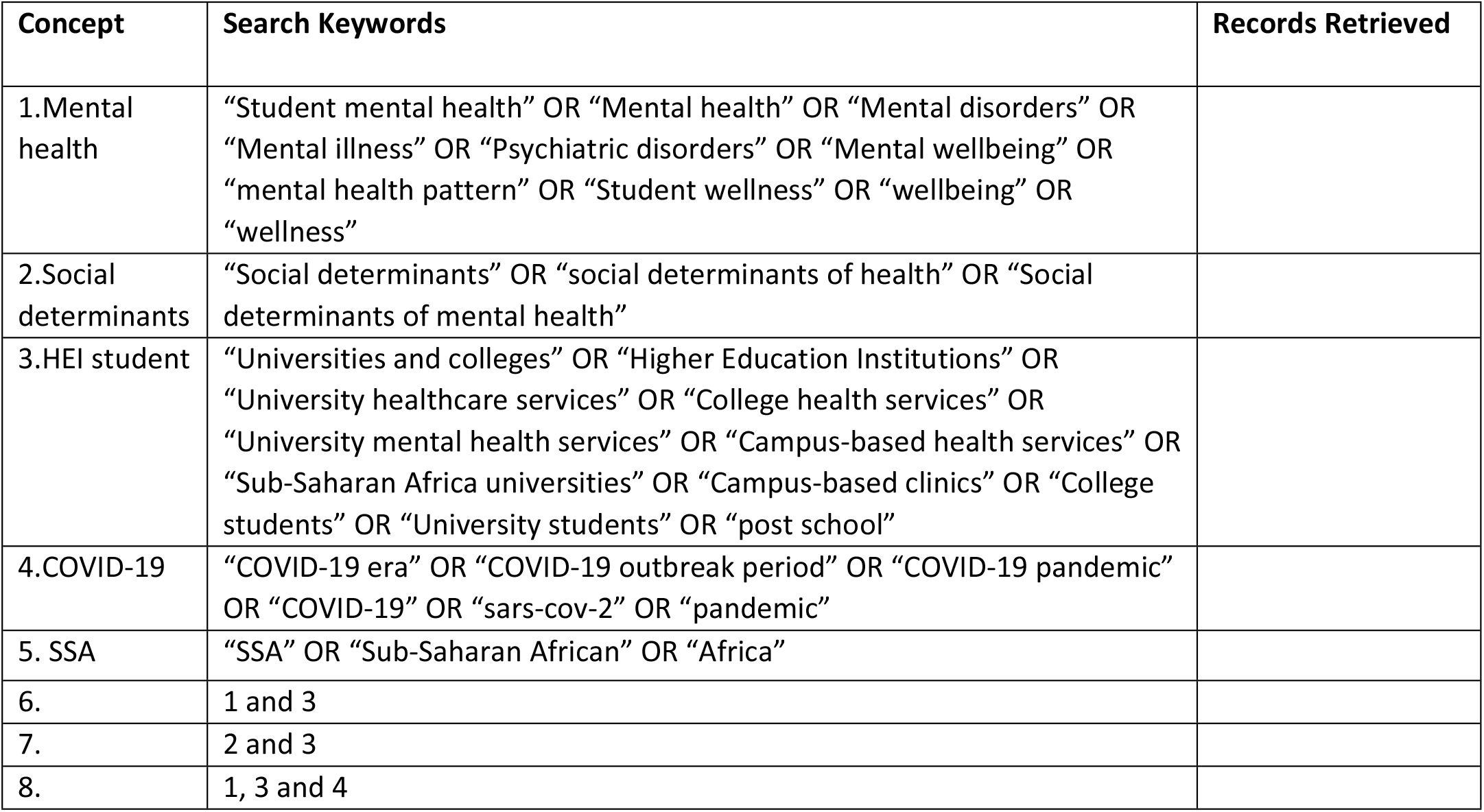
Search.

| Concept | Search Keywords | Records Retrieved |
| --- | --- | --- |
| 1.Mental health | “Student mental health” OR “Mental health” OR “Mental disorders” OR “Mental illness” OR “Psychiatric disorders” OR “Mental wellbeing” OR “mental health pattern” OR “Student wellness” OR “wellbeing” OR “wellness” |  |
| 2.Social determinants | “Social determinants” OR “social determinants of health” OR “Social determinants of mental health” |  |
| 3.HEI student | “Universities and colleges” OR “Higher Education Institutions” OR “University healthcare services” OR “College health services” OR “University mental health services” OR “Campus-based health services” OR “Sub-Saharan Africa universities” OR “Campus-based clinics” OR “College students” OR “University students” OR “post school” |  |
| 4.COVID-19 | “COVID-19 era” OR “COVID-19 outbreak period” OR “COVID-19 pandemic” OR “COVID-19” OR “sars-cov-2” OR “pandemic” |  |
| 5. SSA | “SSA” OR “Sub-Saharan African” OR “Africa” |  |
| 6. | 1 and 3 |  |
| 7. | 2 and 3 |  |
| 8. | 1, 3 and 4 |  |

### Information Sources

The databases which will be searched are MEDLINE (PubMed), PsycInfo, EMBASE, JBI library, Open Access Journals, African Index Medicus, CINAHL (EBSCOhost), and ProQuest, Google Scholar, the Cochrane Library (Ovid), and Grey Literature (various relevant health related websites).

### Study/Source of Evidence Selection

Following the search, all identified citations will be collated and uploaded into EndNote 21 (Clarivate Analytics, PA, USA) and duplicates will be removed. An initial pilot test will be done, and the titles and abstracts of the documents will then be screened by two reviewers against the inclusion criteria. Sources which meet the inclusion criteria will be retrieved, and their citation details imported into the JBI System for the Unified Management, Assessment and Review of Information (JBI SUMARI) (JBI, Adelaide, Australia) [19]. The selected citations will be reviewed using the inclusion criteria by two independent reviewers. Exclusion of documents at full text that do not meet the inclusion criteria will be recorded and reported in the findings. Any disagreements that arise between the reviewers will be addressed by discussing with an additional reviewer. The results of the search and the study inclusion process will be reported in full in the final scoping review and presented in a Preferred Reporting Items for Systematic Reviews and Meta-analyses extension for scoping review (PRISMA-ScR) flow diagram [20].

### Data Extraction

A data extraction tool has been developed by the researchers and will be used for data extraction. Specific details about the participants, concept, context, study methods and main results relevant to the study will be extracted in line with the research question.

A draft extraction form is attached (Table 3). Modifications (if any) to the data extraction form during data collection will be detailed in the report. Any disagreements that arise between the reviewers will be resolved through discussion, or with an additional reviewer.

**Table 3.**
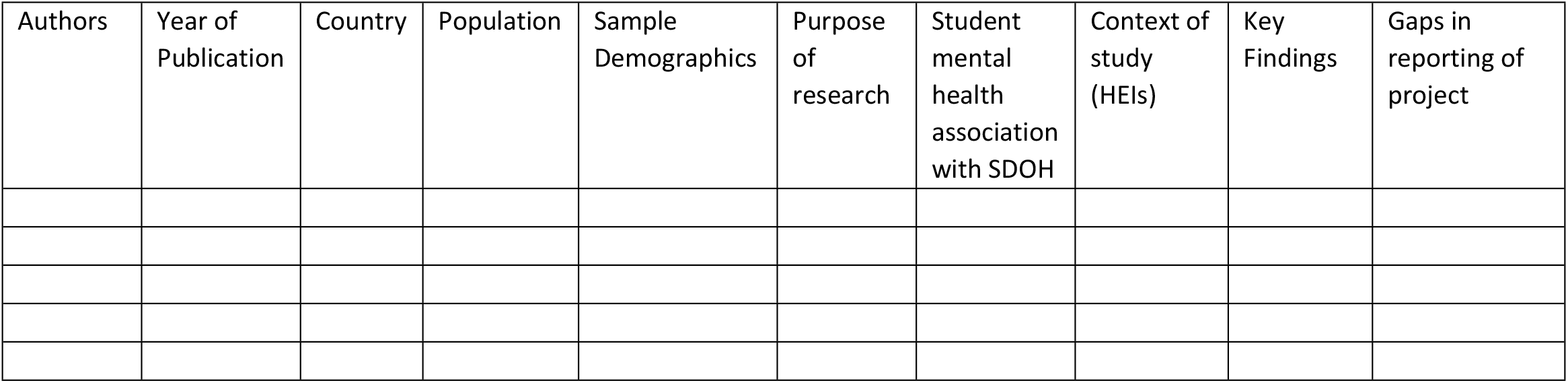
Data Extraction Tool.

### Eligibility criteria

The scoping review will use the participant, concept, context (PCC) approach for eligibility criteria as outlined by [19]. Only documents published during the COVID-19 years 2020 to 2023 from around the SSA region will be included. Documents from outside the COVID-19 period and the SSA geographic region will not be considered. Higher Education Institutions (HEIs) will be post-school universities and colleges, and so school level reports will not be considered.

### Concept

The concept in this scoping review is student mental health during the COVID-19 era by exploring the associated social determinants. This will be the method of collecting, analysing, and presenting data for identifying and mapping student mental health patterns and identifying knowledge gaps in the literature. Studies conducted in Sub-Saharan African for HEIs. This scoping review will provide a SSA context to student mental health and the associated social determinants in HEIs during the COVID-19 era. Documents for data collected outside the SSA region will be excluded as well as publications in languages other than English. Publications before the start of the COVID-19 outbreak period will be excluded.

### Context

This scoping review will include studies from all HEIs where students reported experiencing mental health challenges, diagnosis of mental disorders, accessed university and college mental health services or other student support services, and problems related with student-life. The settings include and not limited to private, public and NGO universities and colleges in SSA. The findings will be used better understand the association between social determinants and student mental health by assessing the extent to which the literature has documented student mental health during the COVID-19 era for universities in SSA.

### Types of Sources

All available peer-reviewed research articles on student mental health in the study period will be included. Both quantitative and qualitative study designs will be included. Analytical observational studies including cohort studies, case-control studies and cross-sectional studies will be considered for the study. Grey literature and opinion papers will also be considered for inclusion.

## Data Analysis and Presentation

After the search, the reviewer will collate and upload all identified documents into EndNote 21. The reviewers will use NVivo -12 to screen titles and abstracts of the documents, and to screen full-text articles and generate themes. A table will be used to extract data and synthesize results and will be reported in accordance with the PRISMA extension for Scoping Reviews (PRISMA-ScR) [20].

## Limitations

A limitation of the study is that only documents published in English will be considered.

### Dissemination

The scoping review findings will be disseminated through a peer-reviewed academic journal, relevant conferences, and to other key experts and stakeholders in the field.

## Data Availability

No datasets were generated or analysed during the current study. All relevant data from this study will be made available upon study completion.

https://doi.org/10.17605/OSF.IO/AXK4Y

## Authors’ Contributions

MM, NM, LC, and SJ: contributed to conception; made significant inputs to planning for the scoping review protocol, revised the manuscript; gave final approval; and agreed to be accountable for all aspects of the work ensuring integrity and accuracy. MM drafted the manuscript.

## Acknowledgements

The authors thank the Dr Norweeta Milburn, University of California Los Angeles for assistance with the data collection tables, Devind Peter University of the Witwatersrand Health Sciences librarian for assistance with initial search strategy guidance.

